# Nurture Early for Optimal Nutrition (NEON) Pilot Randomised Controlled Trial: Qualitative study of community facilitators and attendees’ perspective on intervention delivery

**DOI:** 10.1101/2024.03.09.24304018

**Authors:** Logan Manikam, Priyanka Patil, Ummi Bello, Subarna Chakraborty, Sumire Fujita, Joanna Dwardzweska, Oyinlola Oyebode, Clare H. Llewellyn, Kelley Webb-Martin, Carol Irish, Mfon Archibong, Jenny Gilmour, Phoebe Kalungi, Neha Batura, Rana Conway, Monica Lakhanpaul, Michelle Heys, the NEON Steering Team Membership of the NEON Steering Team is listed in the Acknowledgements

## Abstract

**Background:** Appropriate and healthy feeding practices can enhance a child’s health, prevent obesity, and reduce chronic metabolic disease risks. Given the ethnic variations in feeding practices and metabolic risk, interventions must be community specific. Culturally tailored, grassroots interventions targeting infant feeding can induce behavioural changes, mitigating chronic metabolic disease risks in later life.

**Aim:** The aim of this study was to explore participant feedback and inform intervention delivery methods within marginalised communities.

**Methods:** A pilot three-arm cluster randomised controlled trial was conducted in London’s Tower Hamlets and Newham boroughs, involving community participatory learning and action groups. The study recruited 186 South Asian (Indian, Bangladeshi, Pakistani, and Sri Lankan) mothers or carers of 0-2-year-old children. Attendees were invited to either face-to-face or online intervention arms, facilitated by trained multilingual community facilitators, offering culturally informed discussions on child nutrition and care practices. Qualitative feedback was collected from attendees and facilitators, with thematic analysis identifying key themes, underscoring intervention fidelity and acceptance.

**Results:** Of the initial attendees, 42 (from the remaining 153 at the study’s conclusion) and 9 community facilitators offered feedback on the intervention’s delivery and suggestions for enhancing community-based interventions’ success. Key findings highlighted the need for a more flexible approach to boost participation and the significance of providing accessible, translated documents and resources.

**Conclusion:** Parenting interventions, particularly for new mothers, should adopt a hybrid design. This would provide attendees with the flexibility to select the delivery method, session timings, and the option to participate at any stage of the intervention.

## Introduction

The initial 1000 days post-conception are crucial for a child’s physical and mental development, with nutrition and dental health significantly influenced by birthweight and weight gain during infancy^1^. Consequently, feeding practices during this period can have lifelong effects on a child’s growth and development^2^. Health and age-appropriate feeding practices are essential for overall health, preventing childhood obesity, and reducing the risk of chronic metabolic conditions^2^.

The South Asian (SA) population in the UK exhibits significant disparities and is among the most disadvantaged compared to other minority groups^3^. Despite the adoption of the World Health Organisation’s (WHO) Infant and Young Children Feeding Guidelines, non-recommended complementary feeding practices persist, particularly within South Asian families^4,7^. This noncompliance is attributed to bicultural issues, low levels of acculturation, and conflicting information^4–7^.

Migrants from low-and middle-income countries (LMICs) to high-income countries (HICs) face an increased risk of metabolic disorders^8^. Therefore, culturally tailored interventions targeting infant feeding practices are critical for improved population health.

Currently, few culturally tailored early-life interventions exist in the UK^9^. The UK’s National Health Service (NHS) offers guidelines for optimal nutrition, care, and dental hygiene practices for children, but these are not tailored to specific cultural practices, resulting in suboptimal uptake. Additionally, due to the NHS 10-year Forward Plan and the NHS Five Year Forward View, which aims to unburden the NHS by emphasising health promotion at the community-level, there is a clear need for low-cost culturally sensitive interventions tailored to specific communities^10^.

Community-based participatory interventions could be an effective solution^11^. Past research in LMICs found Participatory Learning and Action (PLA) group approaches to be cost-effective strategies for improving maternal and newborn health^12–15^. Furthermore, this approach is also recommended by the World Health Organisation (WHO) as an effective low-cost strategy for community mobilisation on maternal and newborn health^16^. The PLA approach involves an iterative process where community facilitators guide attendees through a four-stage cycle of identifying issues, designing solutions, implementing them, and evaluating the results^17^.

The Nurture Early for Optimal Nutrition (NEON) programme, designed to enhance infant feeding, care, and dental hygiene practices in South African minority groups, employs a community-based, participatory approach. The programme comprises two stages: NEON 1, a formative and feasibility study, and NEON 2, an intervention development and a pilot randomised control trial (RCT) in two London boroughs – Tower Hamlets (TH) and Newham (NH). Evaluations of such community-based interventions typically utilise mixed-methods, including survey questionnaires, observational data, and qualitative interviews^18^. The study sought to ascertain participant views on the acceptability, feasibility, and generalisability of the NEON community-based PLA group programme and the proposed evaluation methodology for a larger trial.

## Methods

### Study Design

Findings from this study are based on the NEON programme’s 3-arm (face-to-face, online, and control) pilot feasibility cluster RCT in the East London boroughs of TH and Newham NH. Full details of the RCT methodology can be found here^19^.

### Study Population

The target population for this study was SA groups living in London Boroughs of TH and NH.

### Participants

All attendees and CFs who participated in the pilot study were eligible for the qualitative study.

### Outcomes

The following qualitative process measurements were collected to assess the feasibility of proceeding to a definitive trial:

#### Facilitators’ feedback

The PLA group facilitator report form, encompassing both closed and open-ended queries on intervention delivery, participant engagement, fidelity, and the facilitator’s session insights, was collected by the RA from the CFs post each PLA session. Additionally, facilitators underwent interviews, either in-person or telephonically, with a researcher at the six-month follow-up after the final PLA session.

#### Attendees’ feedback

Post the final women’s group PLA session, attendees feedbacked via a digital questionnaire, albeit with low response rates, leading to subsequent invitations for either in-person or telephonic interviews conducted by an independent observer. These feedback mechanisms aimed to identify areas of improvement and address issues pertaining to recruitment, retention, acceptability, fidelity, reach (diversity), measures, and contamination, in line with the topic guides established in NEON 1.

### Analysis

All interviews were digitally audio recorded and transcribed for analysis using the five steps of Framework analysis by Braun and Clarke (supplemental material 1)^20,21^. This study utilized NVivo 12 Plus for a rigorous qualitative analysis of transcripts from CFs and attendees, with a validated coding framework applied to all transcripts. Independent text coding and data analysis by a research assistant and interns identified data-driven themes and concepts, with discrepancies resolved through NEON Steering team discussions. Thematic analysis identified patterns in feedback, using both inductive and deductive approaches in the data analysis process. Interviews, designed within implementation science frameworks, probed key factors such as adaptability, complexity, and compatibility, facilitating the collection of contextual insights and stakeholder perspectives.

## Results

The researchers conducted 9 interviews with CFs (n%=90.00%; n=2 face-to-face and n=7 by phone) and 42 interviews with attendees (n%=29.41%; n=18 face-to-face and n= 24 by phone), between February 2023 and April 2023. The CF’s interviews lasted from 20-40 minutes with a mean duration of 35 minutes whereas the attendees’ interviews lasted for 10-20 minutes with a mean duration of 15 minutes.

From our analysis of feedback/interview data covering the key topics of interest we identified various themes in both facilitators and attendee interviews stated below. These themes and sub-themes are summarised in supplemental materials 2 and 3, are elaborated in detail.

### CF interviews

From interview data, we inductively and deductively identified and explored the following key themes in the facilitators feedback which were further broken down to sub-themes based on responses:

1. Poor attendance of attendees
2. Enhancing attendee engagement
3. Flexibility in session delivery and timings
4. Tool kit
5. Weather conditions
6. Lost attendees
7. Training delivered
8. Support provided by RA
9. Impact on personal growth and community
10. Language and cultural adaptation
11. Data collection challenges
12. Health visitors support
13. Intervention fidelity

#### Attendance & Engagement

##### Poor/Non-Attendance of Attendees (Lack of study knowledge, weather conditions, lost interest, circumstantial unavailability)

The study noted an initial decline in participants, largely attributed to facilitators’ inadequate comprehension of the study’s objectives, impeding their ability to communicate its benefits effectively. Adverse weather conditions, health-related issues among attendees or their children, and changes in personal circumstances like job commencement or relocation further contributed to irregular session attendance.

##### CF dropout (demotivation, waste of effort)

The facilitators’ substantial efforts in participant recruitment and session preparation were compromised by non-attendance, causing demotivation and potential withdrawal, while a lack of understanding of the study’s design and objectives, coupled with the workload, led to attrition among some facilitators.

##### Enhancing Attendee Engagement (Repeated phone calls, WhatsApp reminders, reiterating study benefits)

All facilitators stated that they contacted each attendee multiple times through phone calls and WhatsApp group messages, reminding them of session timings and repeating study benefits to increase attendance and engagement.

#### Intervention Delivery and Fidelity

##### Hybrid model (Flexibility in session delivery and timings [prayer time, etc.])

This was a frequent theme in most facilitator interviews specifying the need to have flexibility in sessions and the opportunity to attend either of the online or face-to-face sessions provided.

##### Tool Kits (Picture cards, translations, updated versions, recipe books)

Everyone reiterated that the tool kit should have been translated, asset map and resource list should have had updated resources available especially considering the cost-of-living crisis. Most facilitators said the recipe book was a hit amongst the attendees and everyone wanted a copy.

##### Training Delivered (Helpful, more sessions needed, involvement)

Many facilitators appreciated the training provided but thought it was rushed over three days and should have been broken down further with more hands-on activities and role plays to boost confidence and preparedness.

##### Intervention Fidelity (Delivered as proposed but this was the problem)

While most facilitators adhered to the manual and delivered the sessions as required, they believed that offering flexibility in timing, duration, and mode of attendance could have enhanced the benefits for the attendees.

#### General Thoughts

##### Support Provided by RA (Helpful, team building)

Facilitators said the RA provided intermittent catchups which helped them share their experiences from sessions with each other, learn from each other, share information, motivate them, and gave them a sense of working in a team helping in team building.

##### Impact on personal growth and community

The facilitators’ involvement in the program, characterized by personal growth and significant community impact through infant nutrition education, is anticipated to result in lasting benefits such as improved child health and parenting skills, highlighting the program’s multifaceted contributions.

##### Language and cultural adaptation

Facilitators’ experiences transcended simple knowledge dissemination, offering personal growth and community benefits, underscored the importance of multilingual resources and cultural sensitivity in areas like child nutrition, and despite language translation challenges, they effectively employed cultural competency training and relevant analogies to enhance understanding.

##### Support Provided by Health Visitors, PPIE (baseline data collection challenges in NH)

In TH, facilitators valued the assistance from HVs and children’s centres for initial data gathering on height and weight, yet encountered difficulties at the second data collection stage due to the requirement for attendees to schedule their own appointments instead of direct outreach from health visitors. Conversely, NH, despite not offering support for the baseline due to time restrictions, effectively handled data collection for the subsequent two timepoints without complications.

##### Data Collection (Very Stressful, useless, poor response)

Facilitators unanimously reported that the survey’s completion was impeded by the use of email links, absence of language translation, and lack of incentives, resulting in a poor response rate despite repeated follow-ups.

##### Findings from sub-themes

###### Low attendee turnout

Initial low turnout led to demotivation among CFs and researchers, impacting the trial and causing some facilitators to resign, but as sessions progressed, increased attendance was observed due to growing community interest and word-of-mouth recommendations.

###### Attendee engagement

CFs utilised regular, personalised communication methods, resulting in enhanced attendee engagement, rapport establishment, and maintained attendance with varying participant engagement levels across sessions.

###### Flexibility in sessions

Attendees’ dissatisfaction with the session structure led to advocacy for a hybrid delivery model, while facilitators’ flexibility in adjusting session timings was crucial for caregivers managing multiple responsibilities.

###### Lost attendees

Facilitators’ limited understanding of RCTs and inability to adequately convey the study’s purpose and requirements were identified as reasons for the initial high dropout rate and attendees’ inability to select their preferred attendance mode.

**Figure 1.**
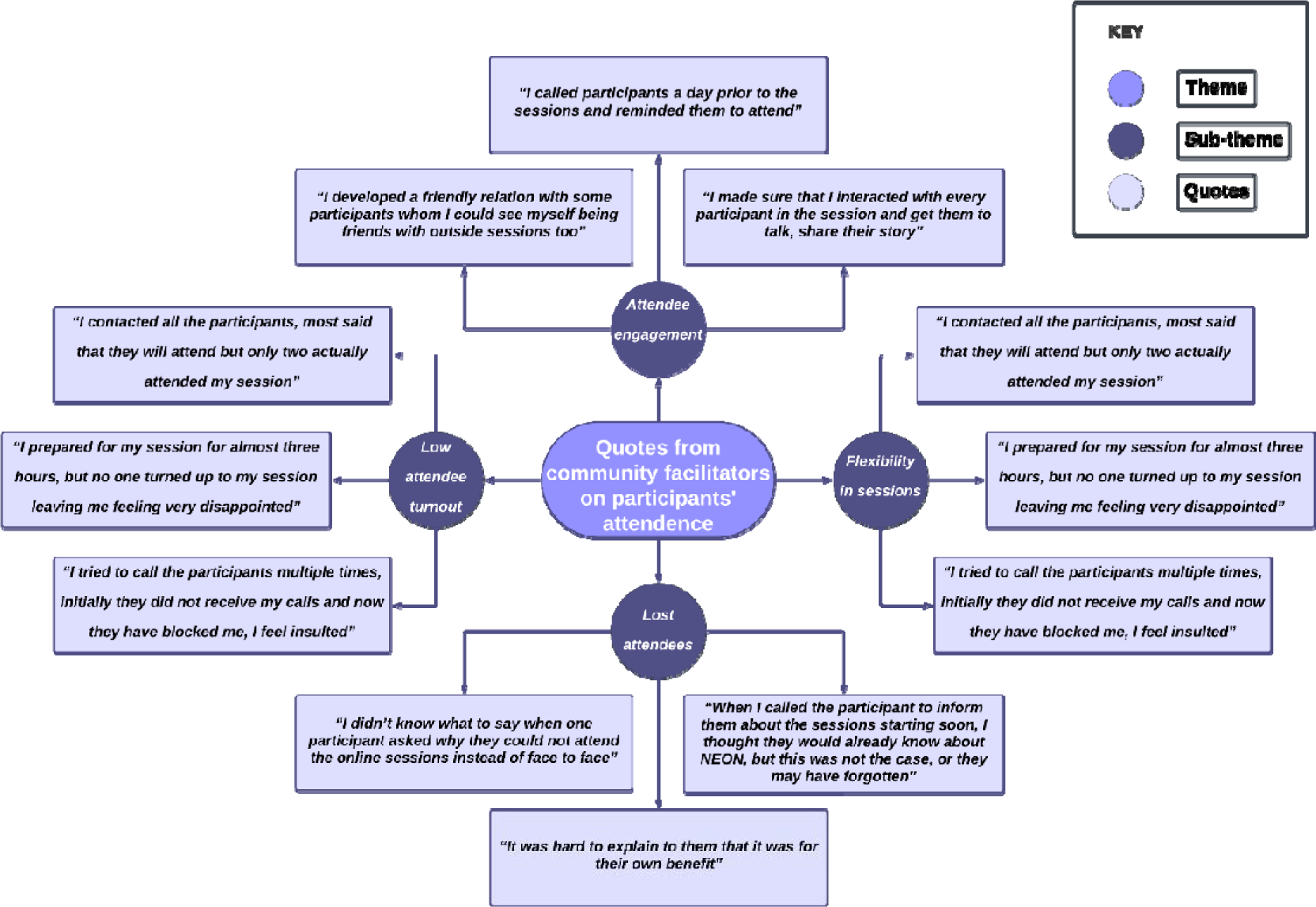
Quotes from CFs on participants’ attendance.

###### Tool kit

Most facilitators, especially those from TH, recommended translating session picture cards into local languages for improved comprehension and suggested revisions to the asset map and resource list to include additional resources, given the cost-of-living crisis. While the recipe book was unanimously deemed the most effective tool for session delivery, adherence to the manual proved challenging for some facilitators, leading them to employ their preferred methods.

###### Weather conditions

Harsh weather conditions in December 2022 and the subsequent care for sick babies significantly impacted both face-to-face and online attendance rates, as noted by several facilitators.

###### Support provided by the RA

Most facilitators said that it was good to have a single point of contact with whom they could discuss any ongoing issues. They suggested their biweekly meetings with the RA helped in team building and acted as a platform for them to share their thoughts on the sessions and challenges faced.

###### Data collection

Facilitators identified the email distribution of questionnaires as a significant barrier due to its user-unfriendliness, proposed translations for improved accessibility, and noted that surveys completed with CFs’ assistance resulted in the highest completion rates due to time efficiency and expert guidance.

**Figure 2.**
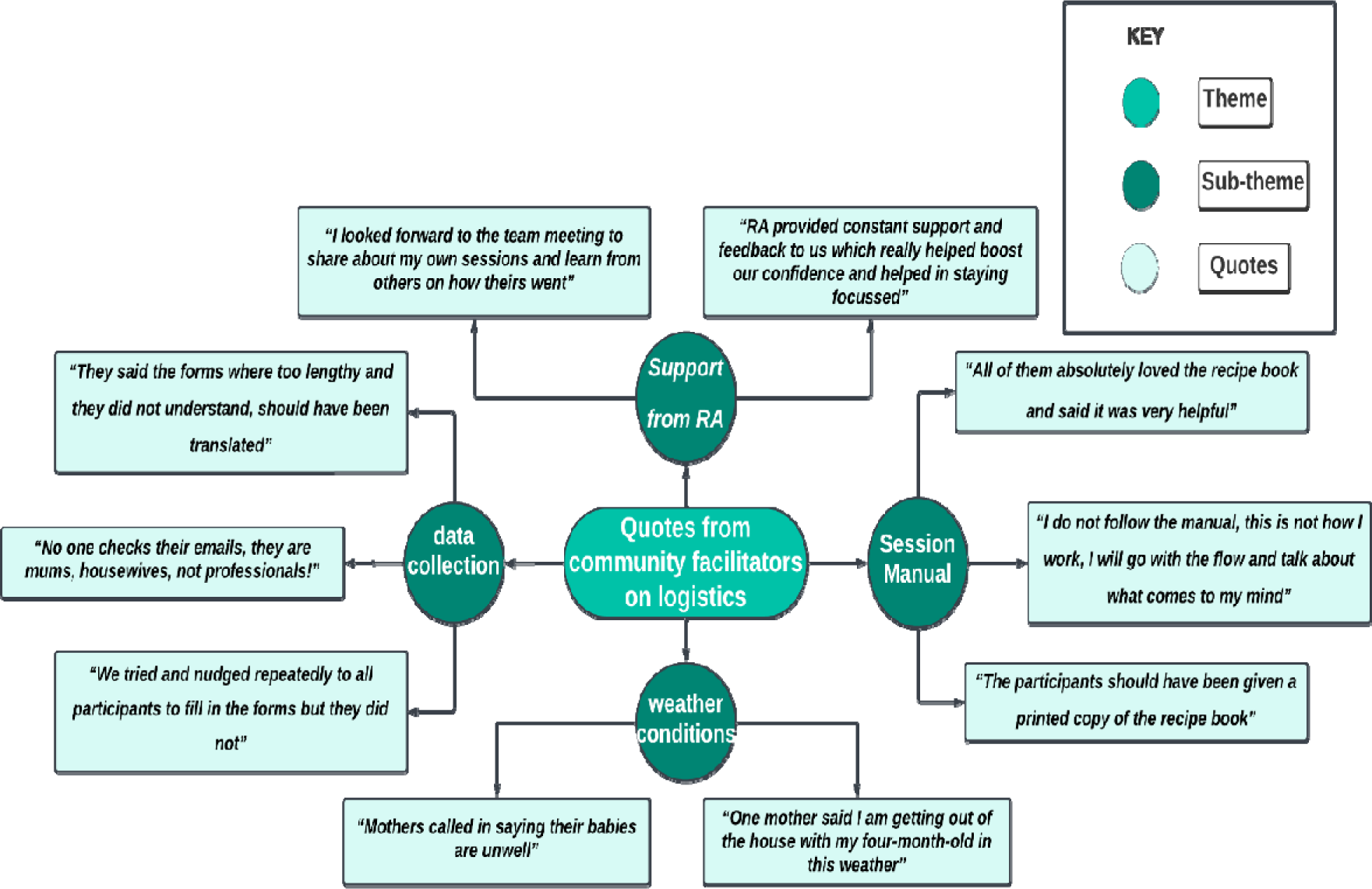
Quotes from CFs on logistics.

###### Training delivery

Facilitators expressed that the three-day training course was very intense and needed more time with shorter sessions to go over the topics thoroughly. They suggested that more role-play activities and exercises would’ve been helpful in improving their delivery skills.

###### Impact on Personal Growth and Community

Through the adaptation of materials to local languages and cultures, facilitators not only enhanced cultural understanding and disseminated knowledge on infant nutrition, thereby boosting self-confidence, but also contributed to education, facilitator empowerment, and community development, with anticipated long-term benefits including improved child health and parenting skills.

###### Language and Cultural Adaptation

Facilitators, transcending mere knowledge transmission, fostered personal and community development through local language and cultural adaptations, thereby engendering participant trust and honouring cultural diversity, and despite the complexities in conveying nutritional concepts, they employed culturally relevant analogies, enhancing their cultural proficiency and making information more comprehensible for participants with limited English proficiency.

###### Health Visitors Support

In TH, facilitators received support from HVs and children’s centres for baseline data collection, aligning with study goals, while in NH, despite initial challenges in attendee measurement sharing due to lack of similar support, the health visiting team’s intervention facilitated data collection for the second time point post-PLA sessions.

###### Intervention fidelity

Most facilitators followed the delivery and structure of the sessions as outlined in the manual. However, some facilitators felt that they could have improved engagement and attendance if they had the flexibility to make minor changes, such as allowing attendees to join sessions of their choice (eg face to face or online).

**Figure 3.**
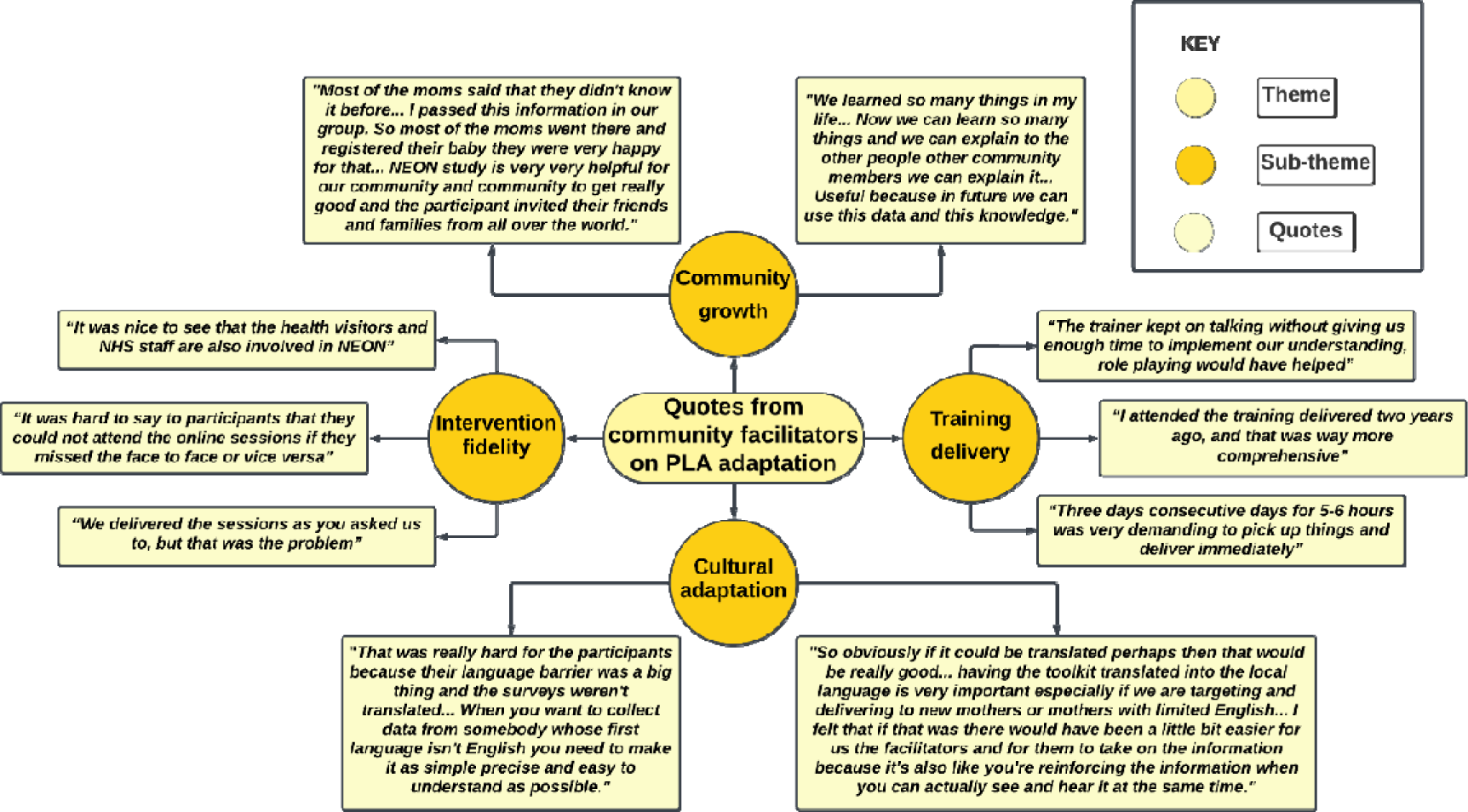
Quotes from CFs on language and cultural adaptation, training, intervention fidelity and co-production.

### Attendees’ interviews

In the thematic analysis of interview data, we identified the following key themes in the attendees’ feedback which are summarised in supplemental material 3.

1. The recruitment process
2. Study explanation
3. Non-participation/attendance
4. Language barrier - picture cards and questionnaire
5. Attitudes to the study
6. Wider reach and inclusion
7. Flexibility in sessions
8. Session benefits

#### Recruitment

Most interested participants were referred by their HVs. However, alterations in personal circumstances impeded some from continuing, and those who became pregnant during the study were excluded due to the study’s inclusion/exclusion criteria.

#### Study explanation

While most participants, introduced to the study via HVs, comprehended its objectives, they expressed confusion about its design, feeling deprived of choice. Conversely, participants in TH reported higher recruitment satisfaction, largely attributed to the engagement of advocacy assistants fluent in their native languages (Bangladeshi and Sylheti), enhancing their comfort levels.

#### Non-participation/attendance

During the study’s initial stage, attendee numbers significantly dropped due to CFs’ contact regarding programme details, personal circumstance changes, and feelings of exclusion, particularly among control arm participants.

#### Language barrier

Most attendees from TH expressed that they would have benefited significantly if the materials shared had been translated into their native languages. However, in NH, the response was more varied, indicating differences in English literacy levels within the two boroughs.

**Figure 4.**
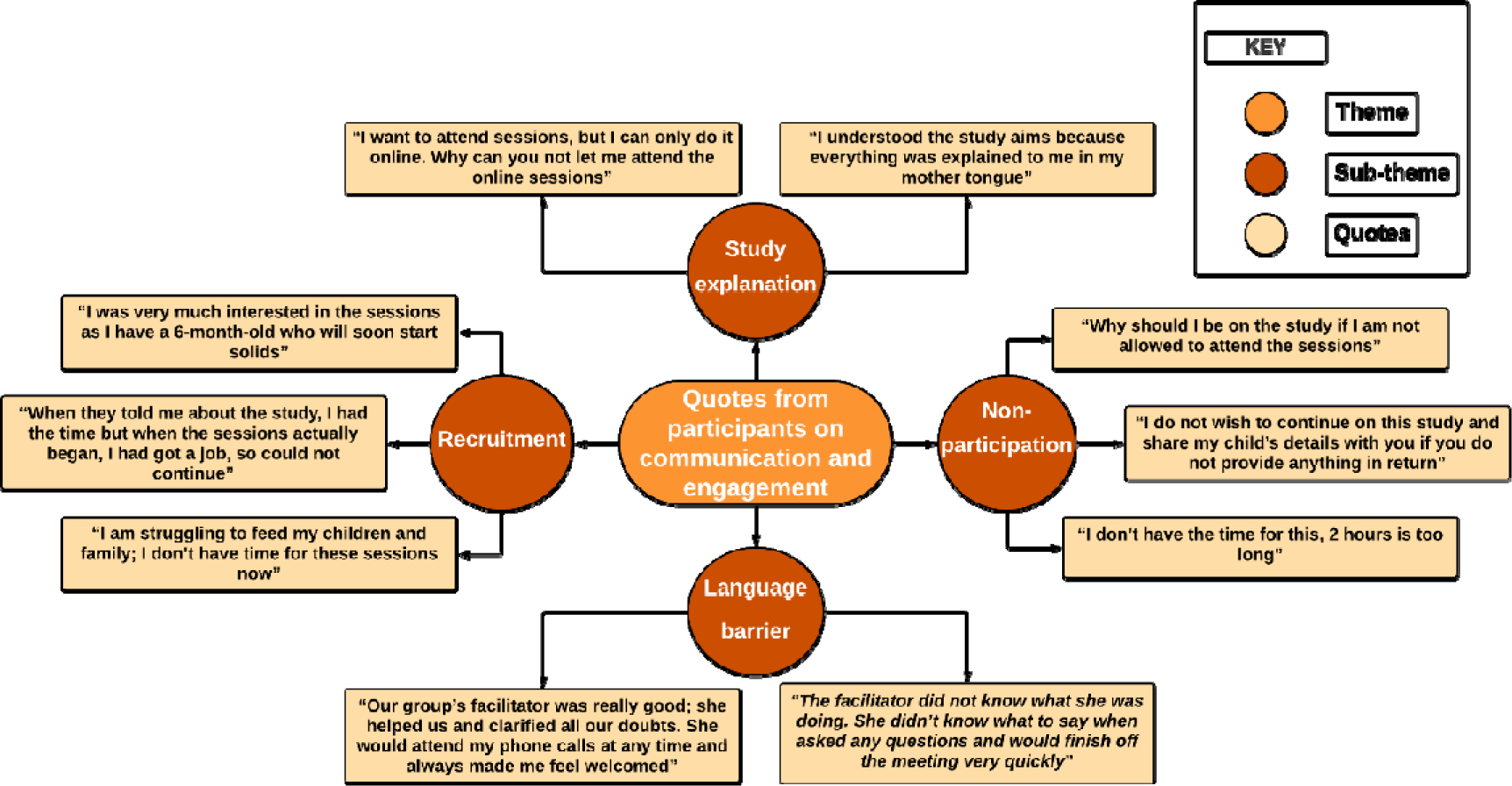
Quotes from participants on language barriers, understanding, recruitment and non-participation affecting communication and engagement.

#### Attitudes to the study

In our study, ‘attitude’ was delineated as any belief, positive or negative, regarding the program’s effectiveness, feasibility, and the perception of the CFs. Our findings indicated that positive attitudes among CFs bolstered attendee commitment and acceptance of the program, while a lack of program awareness and negative CF attitudes adversely impacted attendee trust.

#### Wider reach and inclusion

Regular attendees expressed interest in involving peers with children of similar age, attributing this to the sessions’ informative content, and suggested expanding the program to pregnant women. Parents of 1–2-year-olds regretted their late access to this information, believing it would have influenced initial feeding practices, and recommended introducing these sessions to pregnant women and new mothers to promote early beneficial habits.

#### Flexibility in sessions

Almost all attendees suggested the need for having flexibility in the sessions and allowing them to choose how they wanted to attend these sessions - face-to-face or online.

#### Benefits of the sessions

Participants who attended all eight sessions reported significant learning, which prevented the continuation of suboptimal practices. They expressed a desire for the continuation of these informative and enjoyable sessions, which also fostered a sense of camaraderie and comfort within the group.

**Figure 5.**
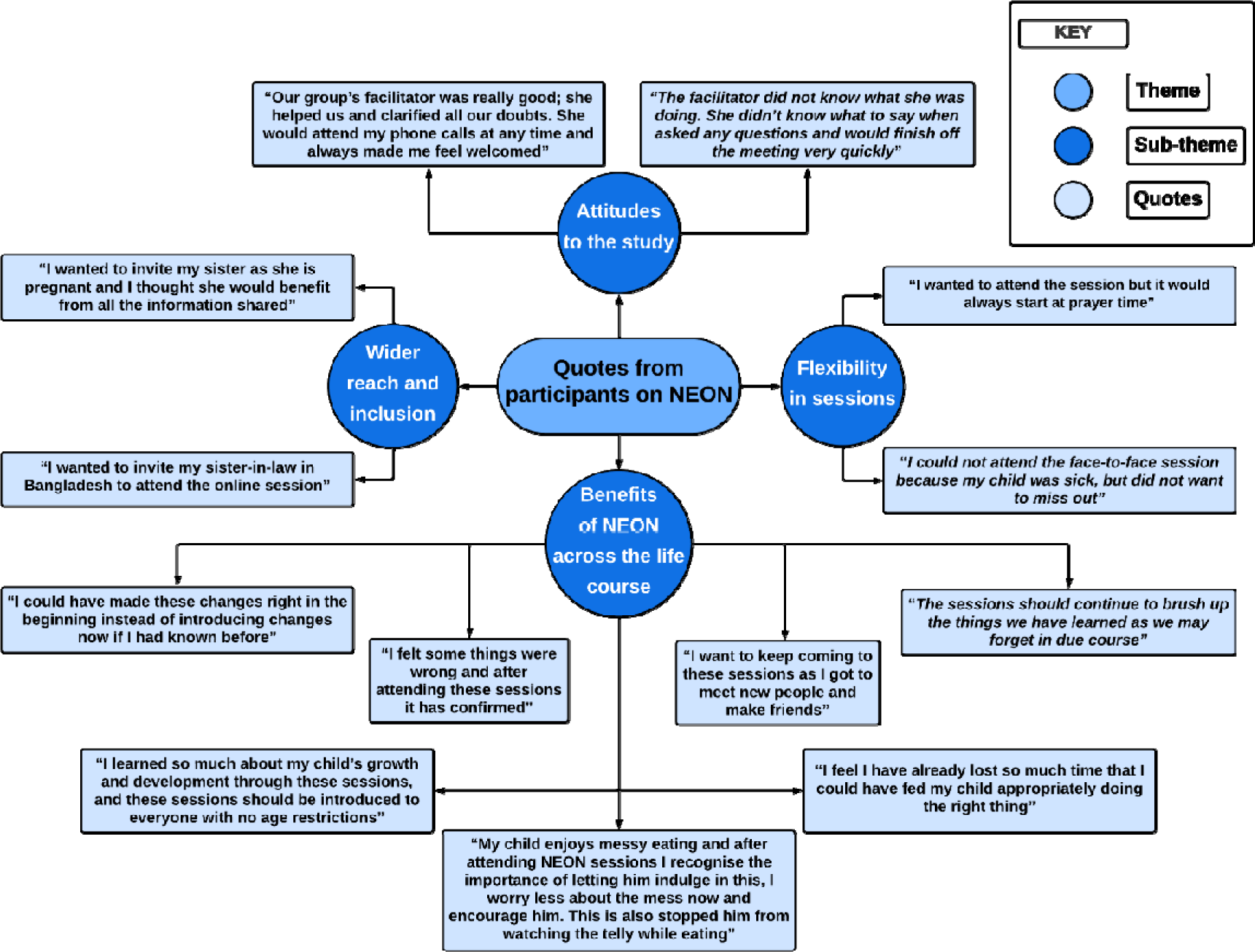
Quotes from participants on NEON.

## Discussion

This study distinctively evaluates the viability of a culturally tailored community PLA group intervention through qualitative data. Participants and facilitators recognized the significance of these structured community interventions in promoting community transformation, advocating for broader initiatives to include larger demographic groups and communities for optimal impact.

### Facilitator Training and Engagement

The facilitators’ comprehension of the study design and objectives was crucial to their engagement. Facilitators who had difficulty understanding the study’s aims and design, despite training, were less successful in representing the study to attendees and addressing their queries. This resulted in subpar session quality, leading to attendee attrition and subsequent facilitator disengagement.

### Mode of Delivery and Intervention Fidelity

Both facilitators and attendees expressed a strong preference for a hybrid delivery model. Despite the RCT having multiple delivery modes, the absence of choice led to early study attrition.

### Resource Utilisation and Data Collection

Both facilitators and attendees found the resources shared during the study sessions highly beneficial. However, most attendees preferred printed and translated versions of the resources, especially the recipe book, over digital copies. This underscores the need for inclusive strategies tailored to specific populations with language barriers.

### Patient and Public Involvement (PPI)

PPI was integrated throughout the study, from the formative phase involving focus group discussions with community members and health professionals, to the intervention development and delivery phase. Facilitators highlighted the support provided by the TH GP care group for outcome measurement, indicating well-organised processes and support.

### Limitations

The study acknowledges potential limitations, including the possibility of social desirability and respondent bias in interviews conducted by CFs. Additionally, some data may have been lost in translation as interviews were not conducted in the primary researcher’s native language.

## Conclusion

Parenting interventions, particularly for new mothers, should adopt a hybrid design. This would provide attendees with the flexibility to select the delivery method, session timings, and the option to participate at any stage of the intervention. to this Conclusion The study suggests a hybrid delivery-model, acknowledging the post-Covid-19 landscape. While digital platforms offer benefits, face-to-face sessions are crucial for community-ties. Enhanced training for CFs, simplified data-tools, and tailored designs are essential for successful community-based interventions.

## Supporting information

Supplementary Material 3: Summary of themes and sub-themes in thematic analysis for Study Attendees

Supplementary Material 2: Summary of themes and sub-themes in thematic analysis for CFs

## Data Availability

All data produced in the present study are available upon reasonable request to the authors.

## Acknowledgements

The authors would like to thank the South Asian community facilitators in the NEON Intervention and community members of the London Boroughs of Tower Hamlet and Newham for their important contribution and engagement to this research project. The authors would like to acknowledge the contribution of the NEON Core Team, Steering Team and all health experts who contributed to this study. We would like to thank the Women & Children First Charity and First Steps Nutrition Trust for their valuable contributions and guidance throughout the study. Steering team members had an opportunity to critically review results and contribute to the process of finalising this paper. The authors would like to thank the National Institute of Health Research (NIHR) Academy and the NIHR Collaboration for Leadership in Applied Health Research and Care North Thames for funding the NEON study. This work is also supported by the NIHR GOSH BRC. The views expressed are those of the author(s) and not necessarily those of the NHS, the NIHR or the Department of Health.

## Collaborators

In addition to the authors, members of the NEON steering team consist of Prof Atul Singhal, Prof Mitch Blair, Joanna Drazdzewska, Dr Sonia Ahmed, Amelie Gonguet, Gary Wooten, Dr Ian Warwick, Vaikuntanath Kakarla, Phoebe Kalungi, Prof Richard Watt, Prof Audrey Prost, Dr Edward Fottrell, Ashlee Teakle, Prof Oyinlola Oyebode, Keri McCrickerd, Dr Rana Conway, Professor Lisa Dikomitis, Mari Toomse-Smith, Scott Elliot, Julia Thomas, Aeilish Geldenhuys, Chris Gedge, Kristin Bash, Dr Dianna Smith, Kate Questa, Dr Megan Blake, Prof Gary Tse, Dr Queenie Law Pui Sze, Gavin Talbot, Dr Chiong Yee Keow, Dr Angela Trude, Prof Lindsay Forbes, Dr Nazanin Zand, Lakmini Shah, Yeqing Zhang, Dina Mobashir, Natasha Chug, Tala El Khatib and Delaney Douglas-Hiley. Contributors ML, LM, MH, NB, CL and PP formed the core team of the study and delivered the research methodology. PP carried out all the activities in the intervention delivery and wrote and devised the paper along with UB, SC and SF. LM, ML, MH, NB, CL, JW, OO, JG, KW-M, CI, MA, and PK validated the study and revised the manuscript critically for important intellectual content. PP, UB and SC contributed to the manuscript writing, and prepared it for submission. LM and PP had primary responsibility for the final content. All authors read and contributed to reviewing the study data, the designing of the manuscript, and the approval of the final manuscript.

## Disclaimer

The views expressed in the publication are those of the author(s) and not necessarily those of the sponsor (UCL), funder (NIHR), study partners (Tower Hamlets GP Care Group, London Borough of Newham Council).

## Patient and public involvement

Patients and/or the public were involved in the design, or conduct, or reporting, or dissemination plans of this research. Refer to the Methods section for further details.

## Research ethics

This study obtained ethical approval from UCL Research Ethics Committee [Ethics ID 17269/001], Sponsor reference number: 142600, Funding Reference: NIHR300020 and IRAS number: 296259, Ref: 21/SW/0142. Written and/or verbal informed consent was obtained from the participants prior to interviews; the participants were ensured about confidentiality and anonymity. All data was protected and made available only to the authors; data was stored in UCL on a secure university server.

## Provenance and Peer Review

Not commissioned; peer reviewed for ethical and funding approval prior to submission.

## Data Sharing Statement

The data supporting the findings of this study is available upon reasonable request from the corresponding author.

## Supplementary Material

**Supplementary Material 1:**
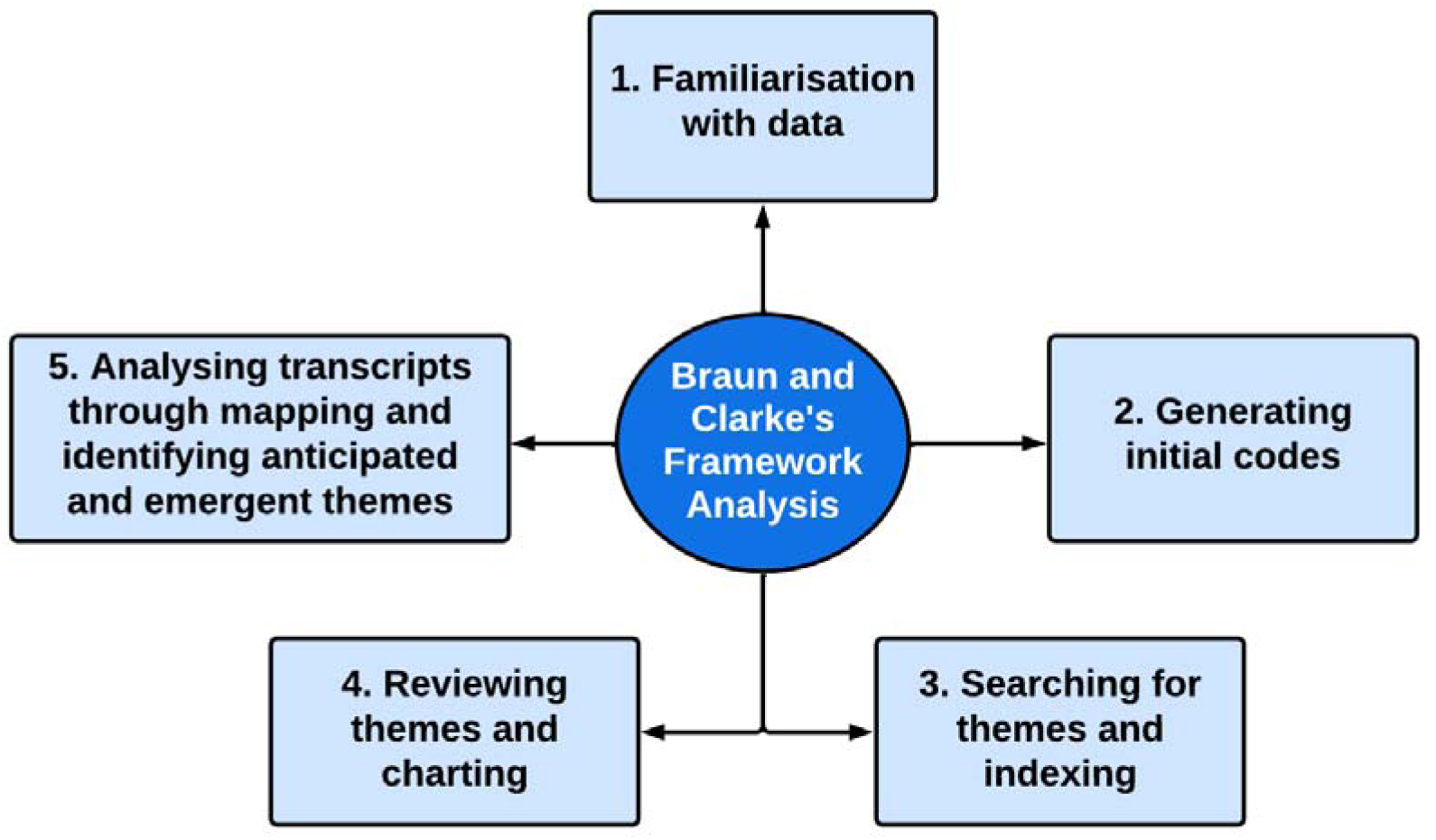
Braun and Clarke’s Framework Analysis.

**Supplementary Material 2:**
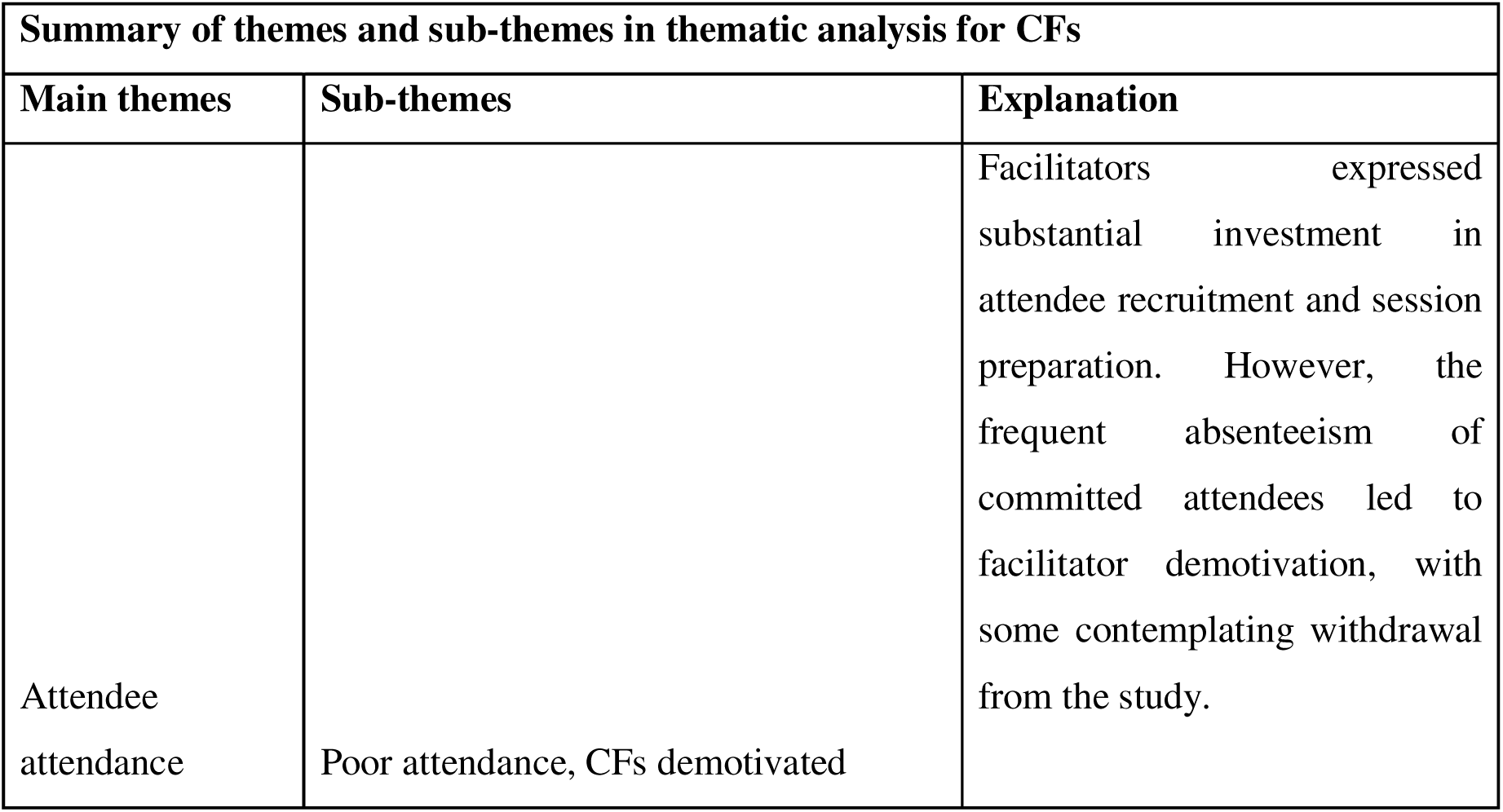
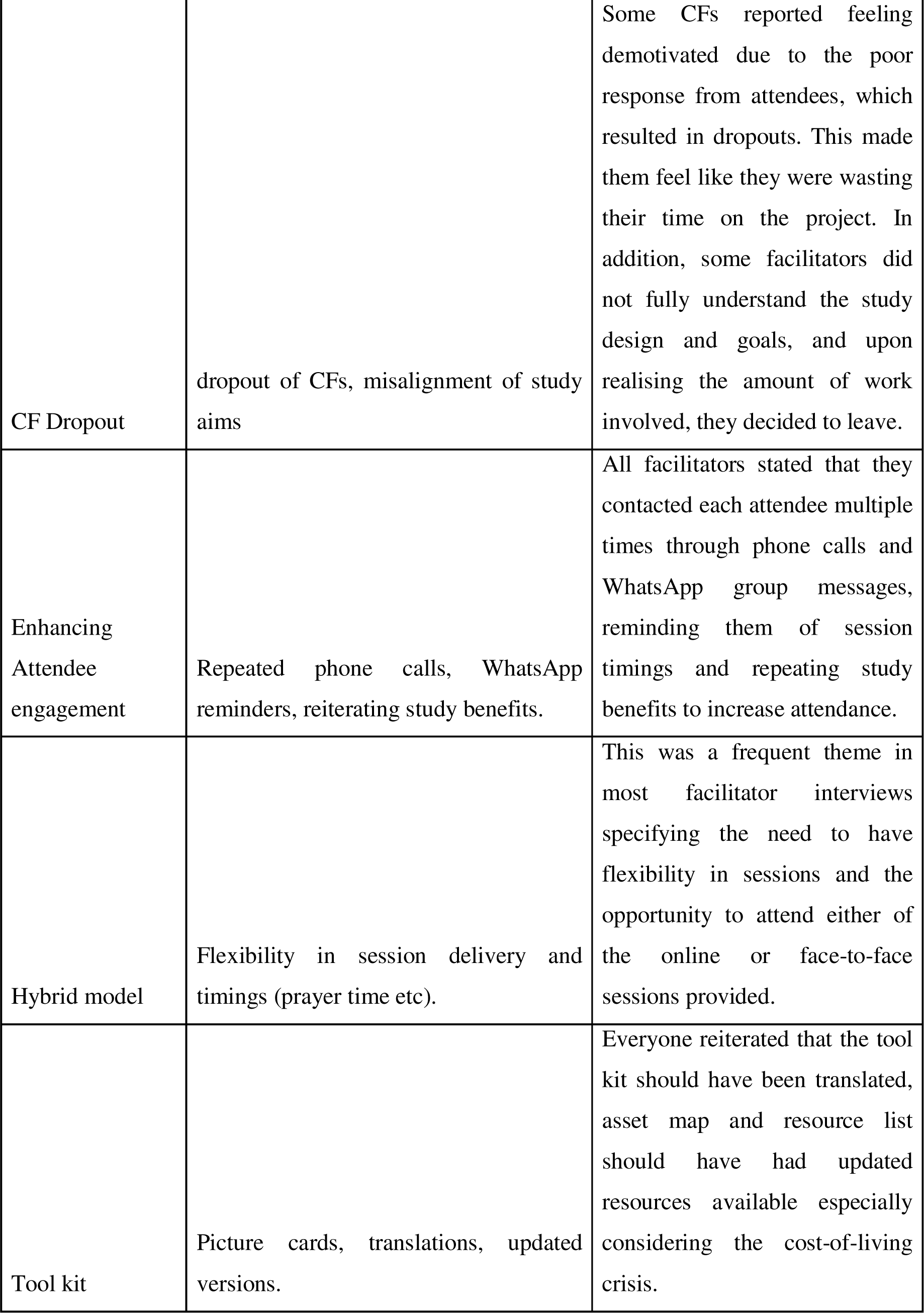
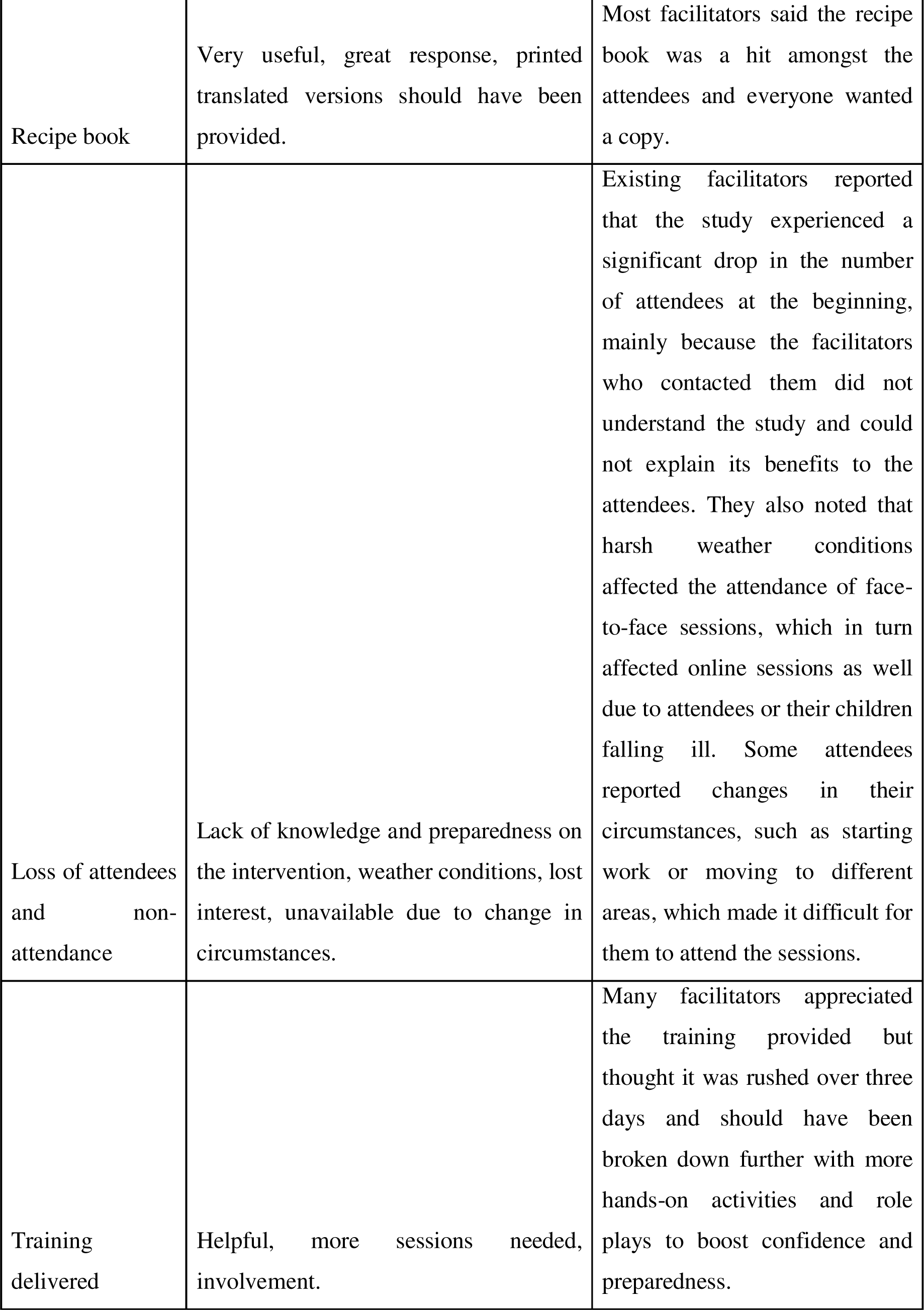
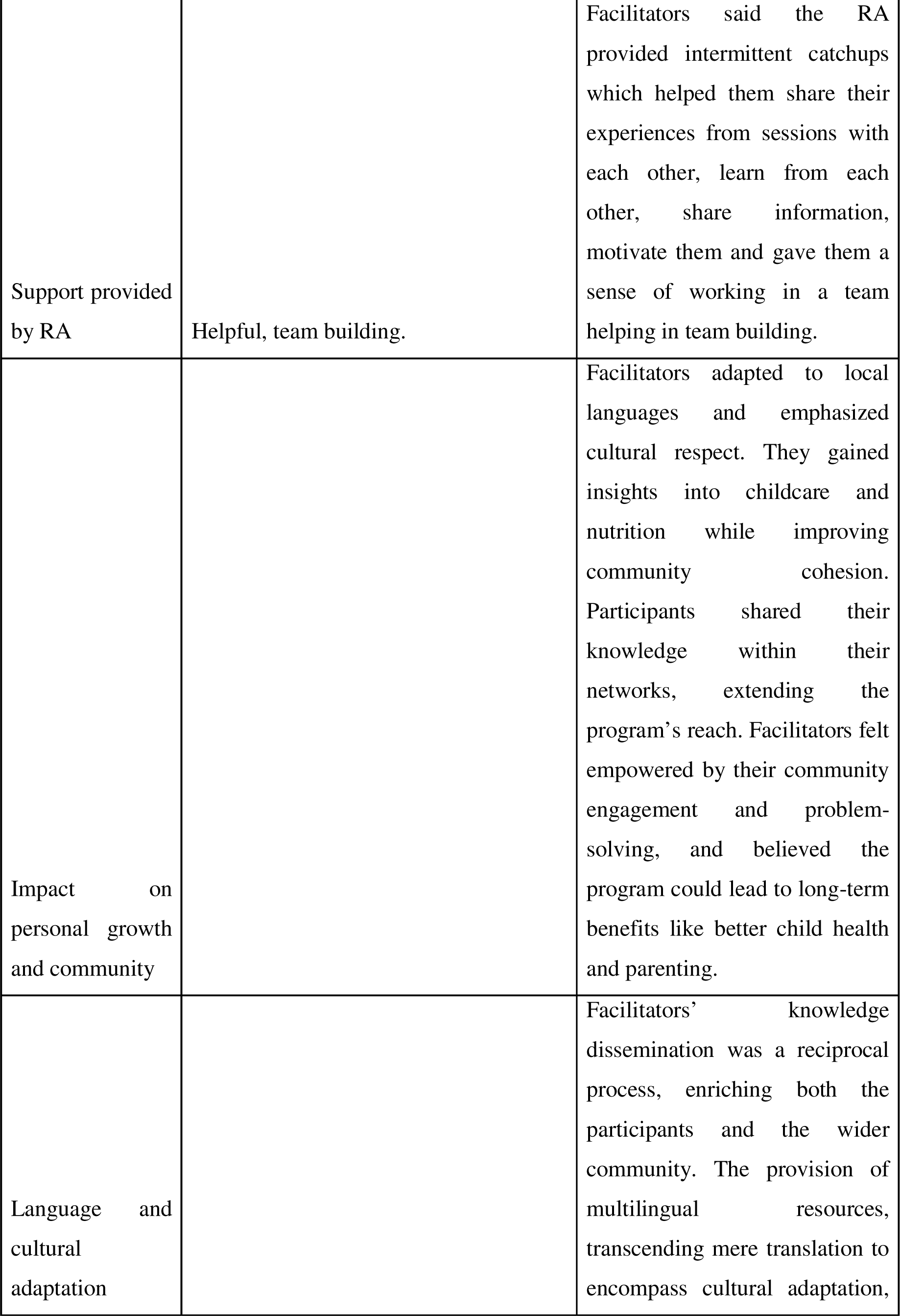
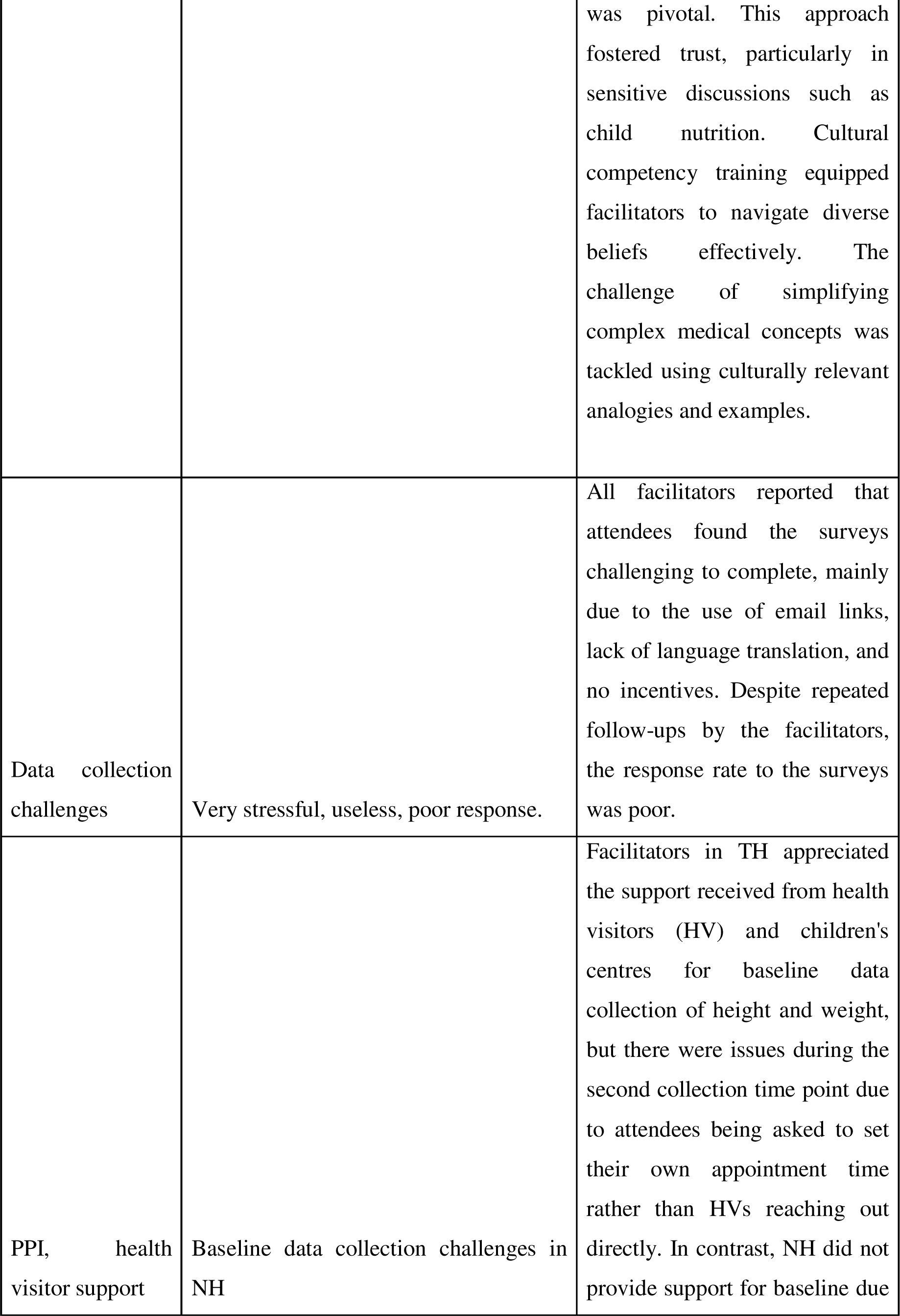
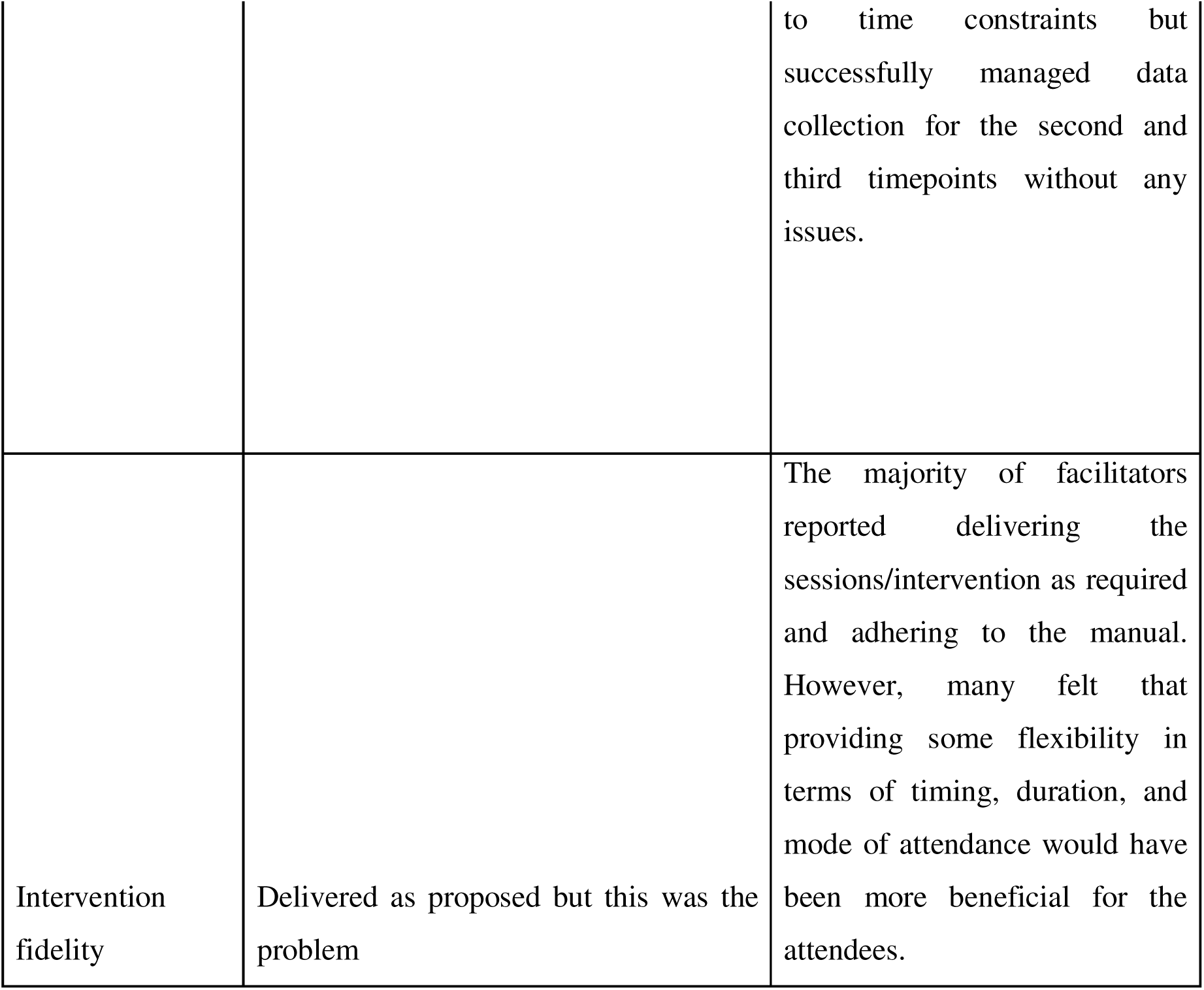
Summary of themes and sub-themes in thematic analysis for CFs.

**Supplementary Material 3:**
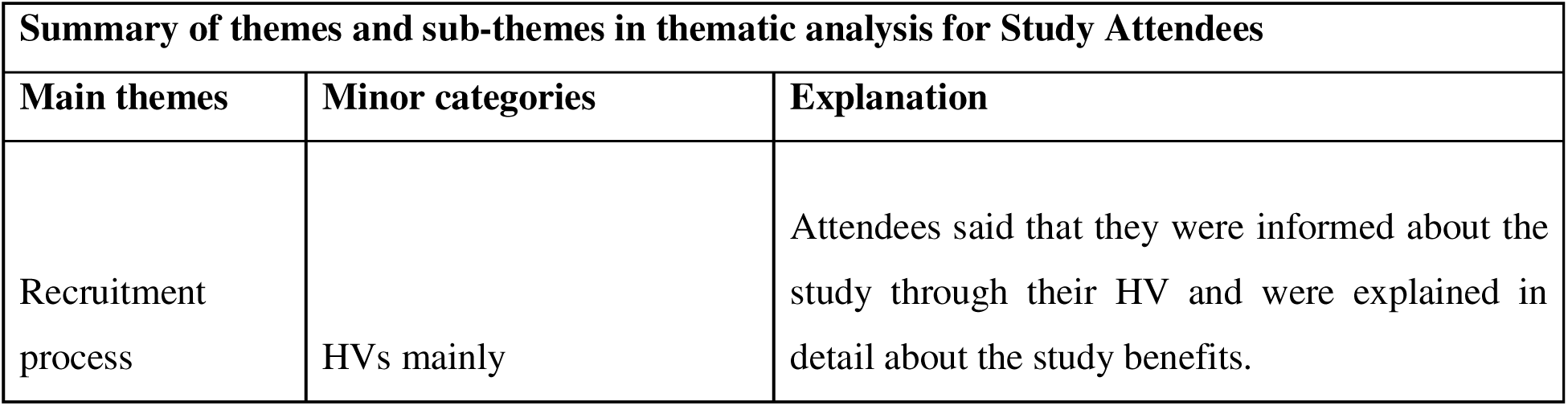
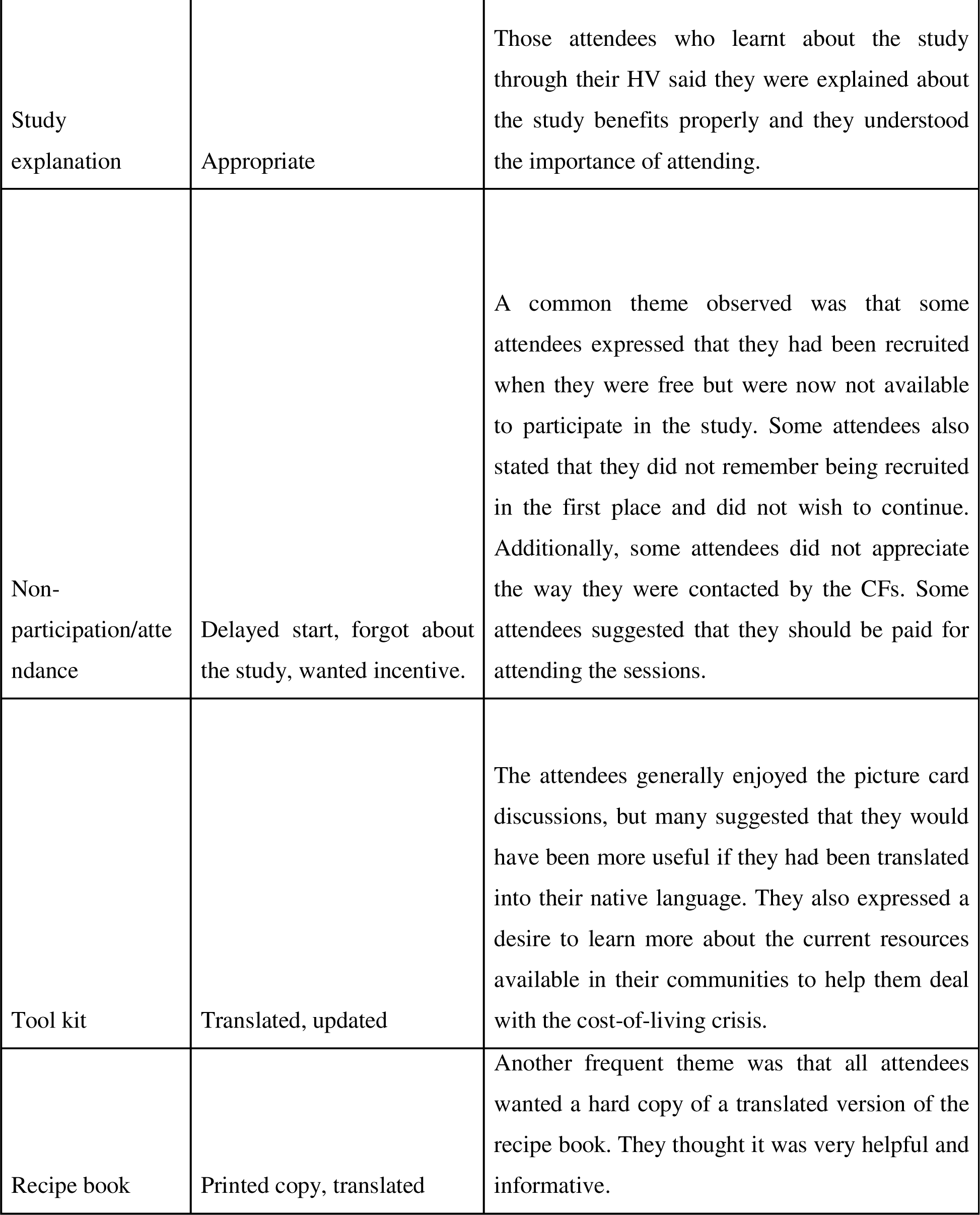
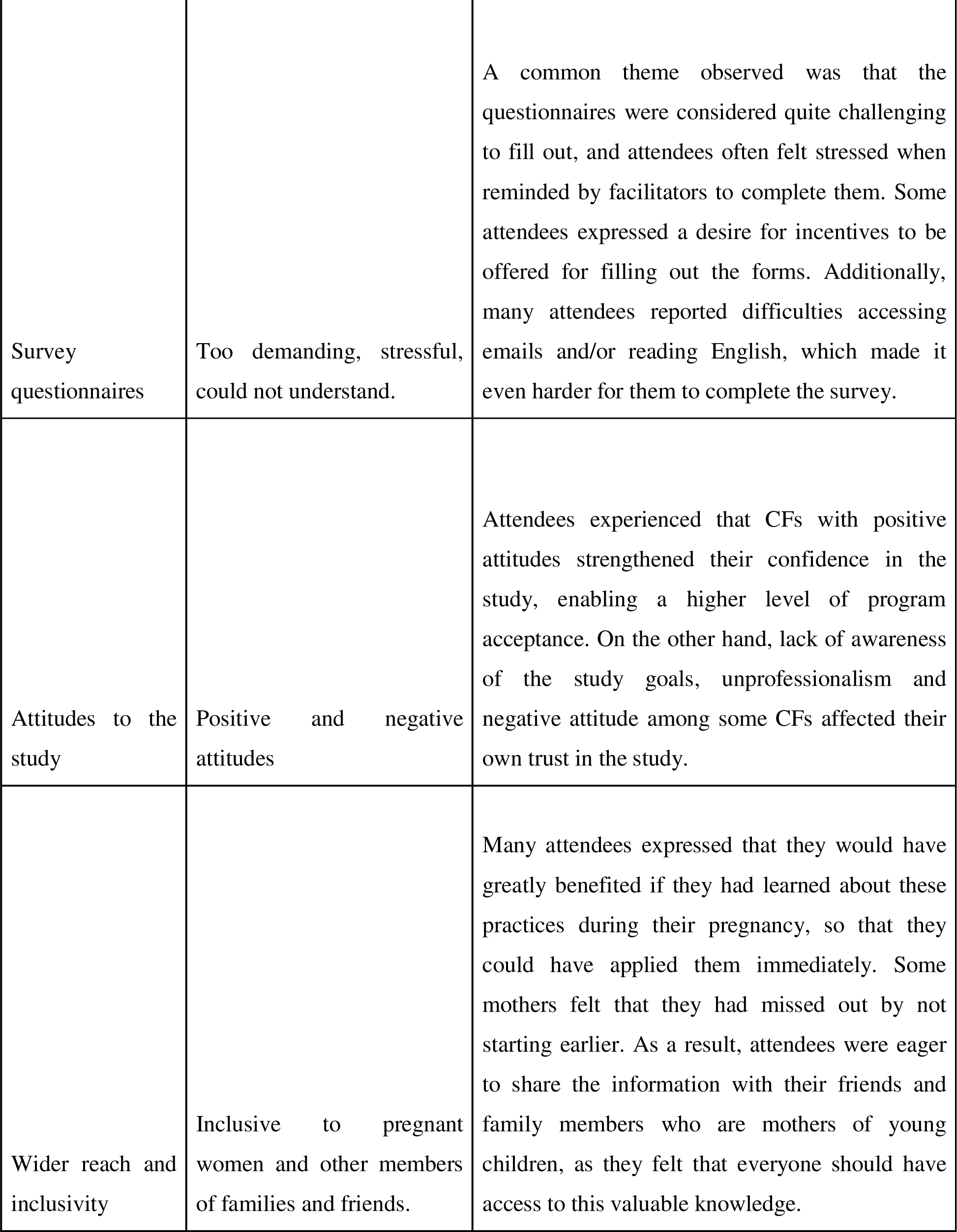
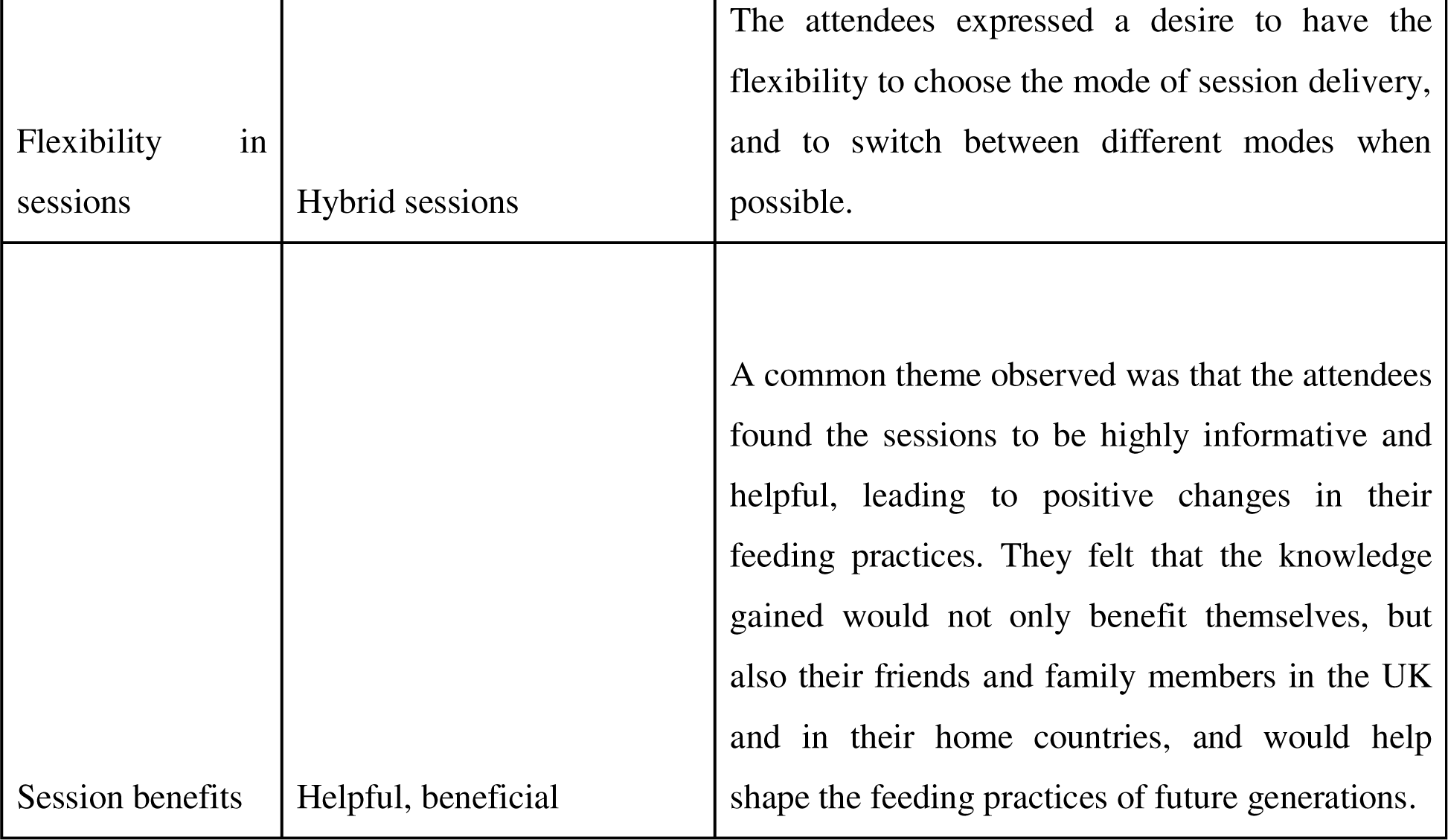
Summary of themes and sub-themes in thematic analysis for Study Attendees.

