## Supplementary Material 3: Summary of themes and sub-themes in thematic analysis for Study Attendees for "Nurture Early for Optimal Nutrition (NEON) Pilot Randomised Controlled Trial: Qualitative study of community facilitators and attendees’ perspective on intervention delivery"

| **Summary of themes and sub-themes in thematic analysis for Study Attendees** | | |
| --- | --- | --- |
| **Main themes** | **Minor categories** | **Explanation** |
| Recruitment process | HVs mainly | Attendees said that they were informed about the study through their HV and were explained in detail about the study benefits. |
| Study explanation | Appropriate | Those attendees who learnt about the study through their HV said they were explained about the study benefits properly and they understood the importance of attending. |
| Non-participation/attendance | Delayed start, forgot about the study, wanted incentive. | A common theme observed was that some attendees expressed that they had been recruited when they were free but were now not available to participate in the study. Some attendees also stated that they did not remember being recruited in the first place and did not wish to continue. Additionally, some attendees did not appreciate the way they were contacted by the CFs. Some attendees suggested that they should be paid for attending the sessions. |
| Tool kit | Translated, updated | The attendees generally enjoyed the picture card discussions, but many suggested that they would have been more useful if they had been translated into their native language. They also expressed a desire to learn more about the current resources available in their communities to help them deal with the cost-of-living crisis. |
| Recipe book | Printed copy, translated | Another frequent theme was that all attendees wanted a hard copy of a translated version of the recipe book. They thought it was very helpful and informative. |
| Survey questionnaires | Too demanding, stressful, could not understand. | A common theme observed was that the questionnaires were considered quite challenging to fill out, and attendees often felt stressed when reminded by facilitators to complete them. Some attendees expressed a desire for incentives to be offered for filling out the forms. Additionally, many attendees reported difficulties accessing emails and/or reading English, which made it even harder for them to complete the survey. |
| Attitudes to the study | Positive and negative attitudes | Attendees experienced that CFs with positive attitudes strengthened their confidence in the study, enabling a higher level of program acceptance. On the other hand, lack of awareness of the study goals, unprofessionalism and negative attitude among some CFs affected their own trust in the study. |
| Wider reach and inclusivity | Inclusive to pregnant women and other members of families and friends. | Many attendees expressed that they would have greatly benefited if they had learned about these practices during their pregnancy, so that they could have applied them immediately. Some mothers felt that they had missed out by not starting earlier. As a result, attendees were eager to share the information with their friends and family members who are mothers of young children, as they felt that everyone should have access to this valuable knowledge. |
| Flexibility in sessions | Hybrid sessions | The attendees expressed a desire to have the flexibility to choose the mode of session delivery, and to switch between different modes when possible. |
| Session benefits | Helpful, beneficial | A common theme observed was that the attendees found the sessions to be highly informative and helpful, leading to positive changes in their feeding practices. They felt that the knowledge gained would not only benefit themselves, but also their friends and family members in the UK and in their home countries, and would help shape the feeding practices of future generations. |
