## Supplementary Material 2: Summary of themes and sub-themes in thematic analysis for CFs for "Nurture Early for Optimal Nutrition (NEON) Pilot Randomised Controlled Trial: Qualitative study of community facilitators and attendees’ perspective on intervention delivery"

| **Summary of themes and sub-themes in thematic analysis for CFs** | | |
| --- | --- | --- |
| **Main themes** | **Sub-themes** | **Explanation** |
| Attendee attendance | Poor attendance, CFs demotivated | Facilitators expressed substantial investment in attendee recruitment and session preparation. However, the frequent absenteeism of committed attendees led to facilitator demotivation, with some contemplating withdrawal from the study. |
| CF Dropout | dropout of CFs, misalignment of study aims | Some CFs reported feeling demotivated due to the poor response from attendees, which resulted in dropouts. This made them feel like they were wasting their time on the project. In addition, some facilitators did not fully understand the study design and goals, and upon realising the amount of work involved, they decided to leave. |
| Enhancing Attendee engagement | Repeated phone calls, WhatsApp reminders, reiterating study benefits. | All facilitators stated that they contacted each attendee multiple times through phone calls and WhatsApp group messages, reminding them of session timings and repeating study benefits to increase attendance. |
| Hybrid model | Flexibility in session delivery and timings (prayer time etc). | This was a frequent theme in most facilitator interviews specifying the need to have flexibility in sessions and the opportunity to attend either of the online or face-to-face sessions provided. |
| Tool kit | Picture cards, translations, updated versions. | Everyone reiterated that the tool kit should have been translated, asset map and resource list should have had updated resources available especially considering the cost-of-living crisis. |
| Recipe book | Very useful, great response, printed translated versions should have been provided. | Most facilitators said the recipe book was a hit amongst the attendees and everyone wanted a copy. |
| Loss of attendees and non-attendance | Lack of knowledge and preparedness on the intervention, weather conditions, lost interest, unavailable due to change in circumstances. | Existing facilitators reported that the study experienced a significant drop in the number of attendees at the beginning, mainly because the facilitators who contacted them did not understand the study and could not explain its benefits to the attendees. They also noted that harsh weather conditions affected the attendance of face-to-face sessions, which in turn affected online sessions as well due to attendees or their children falling ill. Some attendees reported changes in their circumstances, such as starting work or moving to different areas, which made it difficult for them to attend the sessions. |
| Training delivered | Helpful, more sessions needed, involvement. | Many facilitators appreciated the training provided but thought it was rushed over three days and should have been broken down further with more hands-on activities and role plays to boost confidence and preparedness. |
| Support provided by RA | Helpful, team building. | Facilitators said the RA provided intermittent catchups which helped them share their experiences from sessions with each other, learn from each other, share information, motivate them and gave them a sense of working in a team helping in team building. |
| Impact on personal growth and community |  | Facilitators adapted to local languages and emphasized cultural respect. They gained insights into childcare and nutrition while improving community cohesion. Participants shared their knowledge within their networks, extending the program’s reach. Facilitators felt empowered by their community engagement and problem-solving, and believed the program could lead to long-term benefits like better child health and parenting. |
| Language and cultural adaptation |  | Facilitators’ knowledge dissemination was a reciprocal process, enriching both the participants and the wider community. The provision of multilingual resources, transcending mere translation to encompass cultural adaptation, was pivotal. This approach fostered trust, particularly in sensitive discussions such as child nutrition. Cultural competency training equipped facilitators to navigate diverse beliefs effectively. The challenge of simplifying complex medical concepts was tackled using culturally relevant analogies and examples. |
| Data collection challenges | Very stressful, useless, poor response. | All facilitators reported that attendees found the surveys challenging to complete, mainly due to the use of email links, lack of language translation, and no incentives. Despite repeated follow-ups by the facilitators, the response rate to the surveys was poor. |
| PPI, health visitor support | Baseline data collection challenges in NH | Facilitators in TH appreciated the support received from health visitors (HV) and children's centres for baseline data collection of height and weight, but there were issues during the second collection time point due to attendees being asked to set their own appointment time rather than HVs reaching out directly. In contrast, NH did not provide support for baseline due to time constraints but successfully managed data collection for the second and third timepoints without any issues. |
| Intervention fidelity | Delivered as proposed but this was the problem | The majority of facilitators reported delivering the sessions/intervention as required and adhering to the manual. However, many felt that providing some flexibility in terms of timing, duration, and mode of attendance would have been more beneficial for the attendees. |
