## Supplementary figures and images for "Nurture Early for Optimal Nutrition (NEON) Pilot Randomised Controlled Trial: Qualitative study of community facilitators and attendees’ perspective on intervention delivery"

### Supplemental Data 1

**Supplementary Material 1: Braun and Clarke’s Framework Analysis**

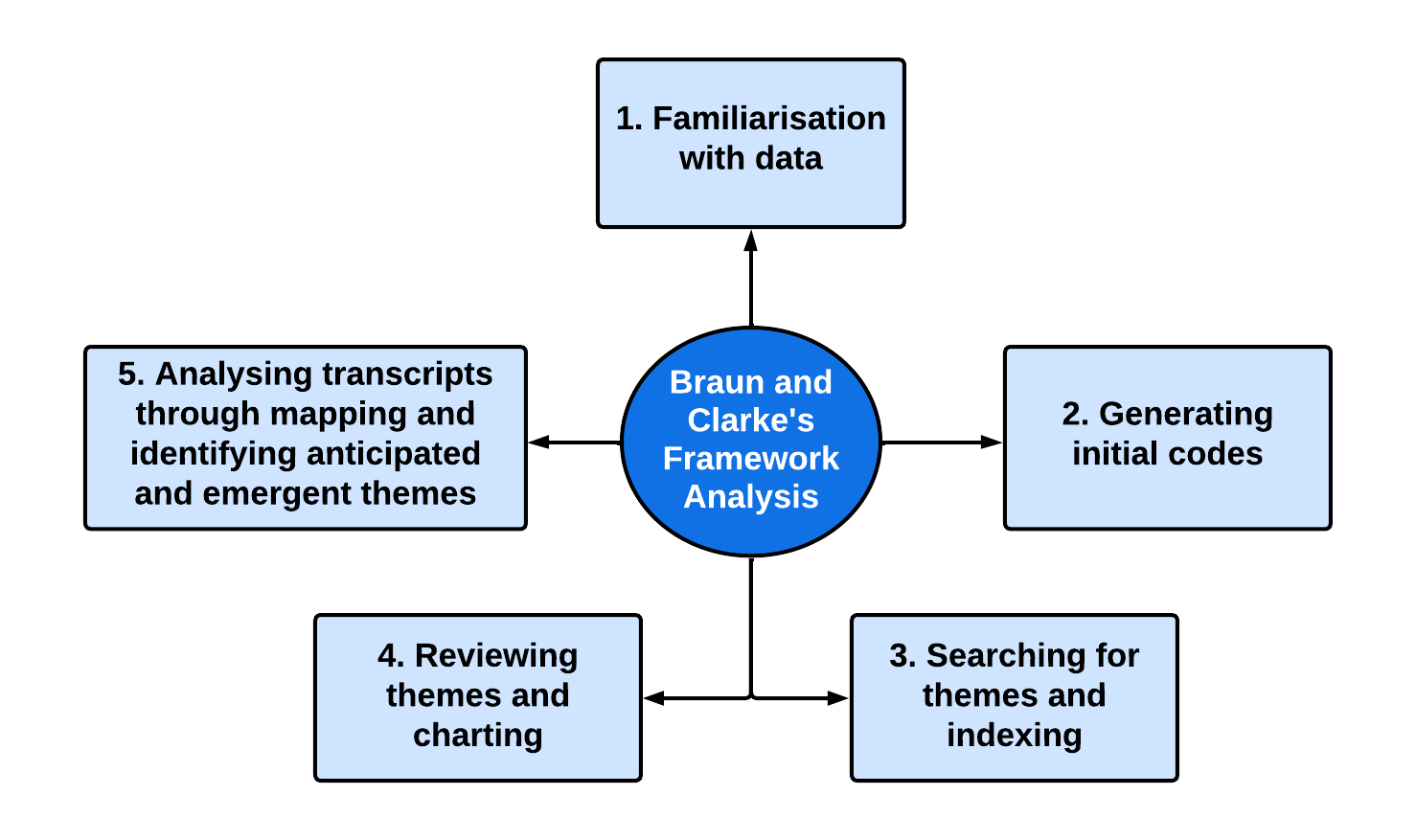
